# Comparative effectiveness of fenfluramine versus cannabidiol in their licensed indications for the treatment of seizures in Dravet Syndrome: a systematic review and network meta-analysis

**DOI:** 10.1101/2022.07.01.22277155

**Authors:** W Linley, M Schwenkglenks, N Hawkins, T Toward

## Abstract

**Purpose:** Fenfluramine and cannabidiol are licensed as add-on therapies for the treatment of seizures in Dravet syndrome (DS); however, there are no comparative trials of these therapies. We assessed the comparative effectiveness of fenfluramine (with/without concomitant stiripentol) versus cannabidiol (with/irrespective of concomitant clobazam, using robust indirect comparison methods.

**Methods:** We systematically searched for randomised controlled trials (RCTs) of licensed regimens published up to 30 November 2021. Outcomes of interest were placebo-adjusted reductions from baseline in monthly convulsive seizure frequency (MCSF), the odds of achieving ≥25%, ≥50%, ≥75% and 100% reductions from baseline in MCSF, and the odds of experiencing serious treatment-emergent adverse events (TEAEs). Comparative efficacy and safety were assessed using Bayesian network meta-analysis (NMA). PROSPERO registration: CRD42021296141.

**Results:** We identified five relevant placebo-controlled RCTs (three for fenfluramine; two for cannabidiol; N=667). All licensed regimens of fenfluramine and cannabidiol significantly reduced MCSF compared with standard of care. When indirectly comparing fenfluramine 0.7mg/kg/day (without concomitant stiripentol) and fenfluramine 0.4mg/kg/day (with concomitant stiripentol) versus cannabidiol 10mg/kg/day, irrespective of clobazam use, the mean differences in placebo-adjusted reduction from baseline in MCSF were 47.3% (95%CrI: 18.9, 64.7) and 35.1% (1.0, 57.5), respectively, and versus cannabidiol 10mg/kg/day plus clobazam were 37.2% (2.0, 59.7) and 23.5% (−20.2, 51.3), respectively. For these outcomes, and for the proportion of patients achieving ≥25%, ≥50% and ≥75% reductions in MCSF, Bayesian treatment ranking indicated ≥99% probability that fenfluramine is the most effective therapy versus <1% probability for cannabidiol 10 or 20mg/kg/day (maximum recommended dose), with/irrespective of concomitant clobazam. Fenfluramine regimens had lower odds of serious TEAEs.

**Conclusion:** NMA using RCT data indicates fenfluramine provides superior convulsive seizure control compared to cannabidiol across all licensed dose regimens and is comparatively well-tolerated. Fenfluramine may meet the need for a highly effective and tolerable add-on therapy to control seizures in DS.

What is already known on this topic

- Fenfluramine (Fintepla®) is a recently licensed add-on therapy to standard of care antiepileptic drugs for the treatment of seizures in Dravet syndrome.
- Cannabidiol (Epidiolex®/Epidyolex®) is also licensed for this use. In Europe cannabidiol is only licensed for use with concomitant clobazam.
- There are no direct comparative data to inform on the relative efficacy and safety of these therapies in the management of seizures in Dravet syndrome. No indirect comparisons have been conducted across the full licensed dose regimens of these therapies.

What this study adds

- Based on NMA using RCT data, fenfluramine provides superior convulsive seizure control in Dravet syndrome compared with cannabidiol across all licensed dose regimens, and is comparatively well tolerated.
- Fenfluramine may meet the need for a highly effective and tolerable add-on therapy to control seizures in Dravet syndrome

## Introduction

Dravet syndrome (DS) is one of the most rare and severe genetic epileptic encephalopathies(1). It typically presents in the first year of life with recurrent convulsive seizures, and progresses to more frequent and more prolonged seizures that may lead to permanent neurological damage (2)[2]. Developmental delay and cognitive impairment are common, exerting a heavy toll on patient and carer activities and quality of life (3,4). Sufferers are at greater risk of premature death compared to the general epilepsy population, due to status epilepticus and sudden unexplained death in epilepsy (SUDEP); 15-20% of children with DS die before reaching adulthood and in those surviving in to adulthood the risk remains elevated(5,6).

Although people with DS may suffer several types of seizures, the most frequent types are tonic-clonic convulsive seizures, which occur in 75-85% of patients(7). High convulsive seizure frequency increases the risk of death, is associated with greater developmental comorbidities and significantly impairs patient and carer quality of life(7,8). A key goal of treatment is therefore to reduce convulsive seizure frequency(9); however, seizures in DS are typically resistant to treatment, including polytherapy with standard of care (SoC) antiepileptic drugs (AEDs), and sustained seizure freedom is rarely achieved(2,10). There is therefore a need for additional therapies that provide meaningful improvements in convulsive seizure control with manageable tolerability.

Cannabidiol (Epidiolex®/Epidyolex®) and, most recently, fenfluramine (Fintepla®) have been licensed in the US and Europe as add-on therapies to SoC AEDs for the treatment of seizures in DS (11–14); however, there are some differences in their labels. Clobazam, a SoC AED used in DS, is a significant treatment effect modifier for cannabidiol and in Europe cannabidiol is only licensed for use with concomitant clobazam. In contrast, clobazam is not a significant treatment effect modifier of fenfluramine (15), making fenfluramine the only add-on therapy for which concomitant clobazam is not specified as a requirement in its label in any jurisdiction. The dose of fenfluramine, however, is determined by whether or not it is added on to AED regimens that include stiripentol (13,14).

Both cannabidiol and fenfluramine were licensed in the US and Europe on the basis of two contemporaneous, similarly designed, placebo-controlled RCTs in which they were added to background SoC AED therapy in patients aged 2-18 years. The primary endpoint in the studies was percentage change from baseline in monthly convulsive seizure frequency (MCSF), assessed over 14–15-week treatment periods. The cannabidiol studies assessed doses titrated to 10 and/or 20mg/kg/day (16,17), and across both doses the placebo-adjusted reduction from baseline in MCSF with cannabidiol was 22-29.8% in the whole trial populations and 36-37% in the subgroup of patients taking concomitant clobazam (18). The fenfluramine studies assessed licensed doses titrated to 0.7mg/kg/day up to a maximum of 26mg/day in patients not receiving concomitant stiripentol [n=119] (Study 1) (19) and 0.4mg/kg/day up to a maximum of 17mg/day in patients receiving concomitant stiripentol [n=87] (Study 2, previously called Study 1504) (20). The placebo-adjusted reduction from baseline in MCSF with fenfluramine 0.7mg/kg/day (without concomitant stiripentol) was 62.3% (19) and with 0.4mg/kg/day (with concomitant stiripentol) was 54.0% (20).

Whilst there is some overlap in the type of adverse events experienced with these add-on therapies (e.g., potential for weight loss, somnolence and lethargy) there are some notable differences in their adverse event risk profiles. Cannabidiol requires periodic liver function testing due to an association with hepatocellular injury, particularly when used with concomitant valproate (11,12), whereas fenfluramine requires periodic echocardiography due to a historical association with valvular heart disease and pulmonary hypertension when used at far higher doses as an anorectic agent for the treatment of obesity in adults (13,14).

It is essential that therapy for DS is tailored to individual needs, taking into account the full risk-benefit profile of therapy options (10). In the absence of direct comparative trials to guide their use, this study was conducted to indirectly compare the efficacy and safety of fenfluramine, as the newest add-on therapy option, against cannabidiol across their licensed dose regimens in DS, using network meta-analysis (NMA).

## Methods

### Literature searches and study selection

Systematic searches for English language published RCTs of fenfluramine and cannabidiol at their licensed doses in DS were initially conducted in PubMed and Embase^®^ up to 28 June 2020 and then updated 30 November 2021 (PROSPERO registration: CRD42021296141). Search strategies included DS-related terms combined with a Cochrane highly sensitive search filter for RCTs (sensitivity and precision maximising version) (21) (see **Supplementary Information Table S1**). Supplementary searches of Cochrane Central Register of Controlled trials (CENTRAL), the Cochrane Database of Systematic Reviews and abstracts from key conference proceedings 2017-21 were conducted using free text DS terms and MeSH headings where available (see **Supplementary Information Table S2**).

Only RCTs reporting outcomes for at least 10 subjects per treatment arm were included. Titles and abstracts were screened by two independent reviewers against the full eligibility criteria listed in **Supplementary Information Table S3**. Data extraction was performed by one reviewer and validated by the other.

The efficacy outcomes of interest were reductions from baseline in monthly convulsive seizure frequency (MCSF), responder rates defined by ≥25%, ≥50% (clinically meaningful), ≥75% (profound) reductions from baseline in MCSF, and convulsive seizure freedom (100% reduction), on the basis that convulsive seizures are the key efficacy endpoint upon which add-on therapies have been licensed (11–14) and are associated with poor outcomes, including seizure related mortality (22–24). Given their individual adverse event risk profiles, safety outcomes of interest were the incidence of any serious treatment-emergent adverse events (TEAEs), and fenfluramine- and cannabidiol-related adverse events of interest.

Feasibility assessment for conducting indirect treatment comparisons (ITCs) for fenfluramine versus cannabidiol were undertaken following the general approach outlined by Cope et al, 2014 (25). Study designs and eligibility criteria, the endpoints used to determine the outcomes of interest and baseline characteristics of patients enrolled in the trials were compared to determine the degree of similarity. Risk of bias was assessed using the Cochrane Risk of Bias 2 tool (26).

### Data synthesis and analysis

As relevant data are available from multiple trials of each intervention, ITCs were conducted using Bayesian NMA where possible. These were performed using MetaInsight v3.14 (August 2020), which employs netmeta and GeMTC packages in R statistical software (27). Fixed effect NMA models were conducted for each outcome given the small number of studies providing data and the lack of repetition of treatment comparisons in the network (**Figure 1**), with alternative models explored in sensitivity analyses. Supplementary Bucher pairwise ITCs (28) of published data were conducted using an online Excel-based tool (29) as a consistency check.

**Figure 1.**
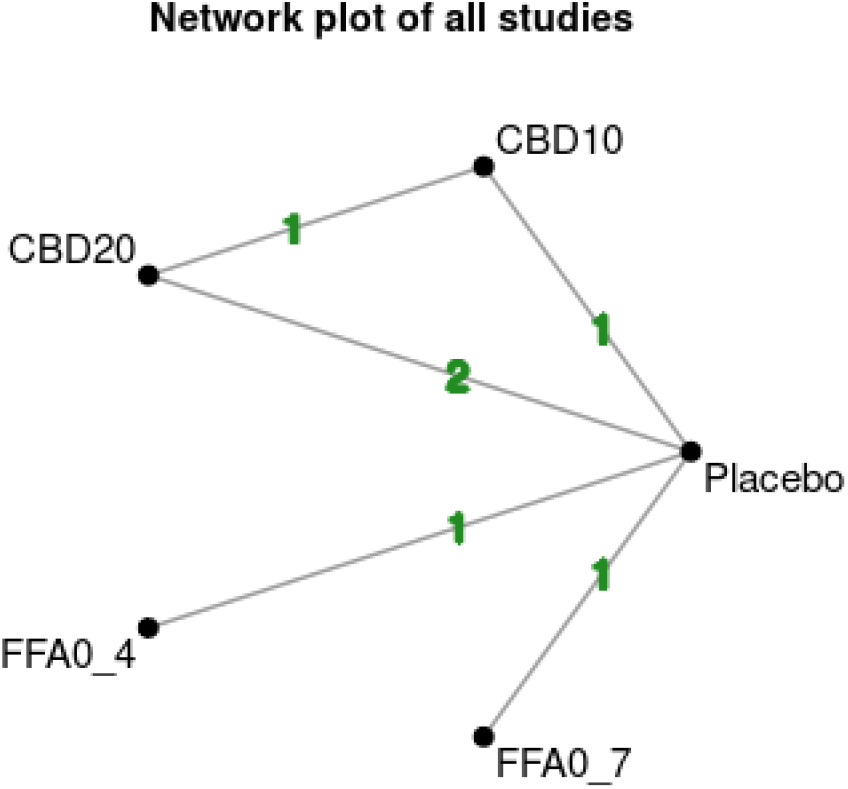
Trial network for all outcomes (full publications) Key: CBD10, cannabidiol 10mg/kg/day; CBD 20. Cannabidiol 20mgmg/kg/day; FFA04, fenfluramine 0.4mg/kg/day up to a max. 17mg/day; FFA07, fenfluramine 0.7mg/kg/day up to a max. 26mg/day. Numbers on lines represent number of trials providing comparison. Excludes fenfluramine Study 3 that at time of literature searches was published as a poster only.

Data for the percentage change from baseline in MCSF were based on parametric analyses of the mean monthly percentage change from baseline compared with placebo reported in the trial publications. These data were log transformed prior to analysis, and the mean differences from the NMAs were back transformed to the ordinary scale. Responder analyses and serious TEAEs were based on data taken without adjustment, and results from these NMAs presented as odds ratios, as used in a previously published NMA in DS (30). Bayesian treatment ranking probabilities were used to indicate the best-to-worst hierarchy of interventions in the trial network. Individual adverse events of interest were considered descriptively.

Analyses versus cannabidiol were conducted by dose in the overall cannabidiol trial populations and, where data allowed, in the subgroup of cannabidiol recipients taking concomitant clobazam, to reflect its licensed indications in different jurisdictions. As clobazam is not a significant treatment effect modifier of fenfluramine (15), data from the overall fenfluramine trial populations were used for all analyses.

The NMAs were reported according to the preferred reporting items for systematic reviews and meta-analyses involving network meta-analysis (PRISMA-NMA) statement (31) (**Supplementary Information Table S4**).

## Results

### Literature search results

The systematic literature searches identified the fully published phase 3 placebo-controlled RCTs that supported the licensing of fenfluramine (Study 1 (19)and Study 2 (20)) and cannabidiol (GWPCARE1B (16) and GWPCARE2 (17)) in DS. An additional phase 3, placebo-controlled fenfluramine RCT (Study 3), published only as a conference poster at the time of the search date cut-off (32), was also identified. No other RCTs fulfilled the inclusion criteria (see PRISMA flow diagram in **Supplementary Information Figure S1**). Feasibility assessment indicated that the trials were broadly similar in terms of designs and assessment of efficacy and safety endpoints over the whole of their 14-15 week treatment periods (**Supplementary Information Table S5**), and the baseline characteristics of recruited participants (**Supplementary Information Table S6**). The fenfluramine and cannabidiol RCTs were at low risk of bias, and although there were some concerns for analyses of the cannabidiol subgroup taking concomitant clobazam (arising from the post hoc nature of those analyses), no domains of the studies were assessed to be at high risk of bias (**Supplementary Figure S2**).

NMAs and Bucher pairwise indirect treatment comparisons were therefore conducted based on efficacy and safety data from the fully published fenfluramine and cannabidiol RCTs (**Figure 1, Supplementary Information Table S8 and S9**). As the third fenfluramine RCT (Study 3) was published only as a conference poster, this was included in NMA sensitivity analyses where data allowed. As the primary publications of the cannabidiol RCTs do not report data for the subgroup of patients taking clobazam, these data were obtained from the European Summary of Product Characteristics (12), a published overview of the cannabidiol trial data (18), and a German Federal Joint Committee (Gemeinsamer Bundesausschuss, G-BA) health technology assessment report (33).

### Mean reduction in monthly convulsive seizure frequency

All licensed regimens of fenfluramine and cannabidiol when added to SoC AEDs significantly reduced the mean MCSF (**Table 1, Supplementary Figure 3**). In NMAs indirectly comparing fenfluramine 0.7mg/kg/day (without concomitant stiripentol) and fenfluramine 0.4mg/kg/day (with concomitant stiripentol) versus cannabidiol at a maintenance dose of 10mg/kg/day, irrespective of clobazam use, the mean differences in placebo-adjusted reduction from baseline in MCSF were 47.3% (95% credible interval [CrI]: 18.9, 64.7), and 35.1% (1.0, 57.5), respectively, in favour of fenfluramine. The mean differences versus cannabidiol 20mg (maximum licensed dose), irrespective of clobazam use, were 50.3% (26.7, 66.4) and 39.3% (9.5, 59.3), respectively, in favour of fenfluramine (**Table 2**).

**Table 1.** Bayesian pairwise placebo-adjusted mean reductions from baseline in MCSF for cannabidiol and fenfluramine licensed dose regimens.

|  | **Cannabidiol irrespective of concomitant use of clobazam (US licensed indication)** |  | **Fenfluramine irrespective of concomitant use of clobazam** |  |
| --- | --- | --- | --- | --- |
|  | CBD10 | CBD20 | FFA0.4 | FFA0.7 |
| Mean difference vs placebo (95%CrI) | 29.2% (8.7, 44.8) | 24.1% (6.8, 38.2) | 54.0% (35.4, 67.0) | 62.3% (47.6, 72.7) |
| Probability Ranked Best | 0.00% | 0.00% | 20.40% | 79.58% |
|  | **Cannabidiol subgroup on concomitant clobazam (EU licensed indication)** |  | **Fenfluramine irrespective of concomitant use of clobazam** |  |
|  | CBD10_CLB | CBD20_CLB | FFA0.4 | FFA0.7 |
| Mean difference vs placebo (95%CrI) | 39.8% (18.6, 55.5) | 36.5% (18.7, 50.3) | 54.1% (35.7, 67.4) | 62.3% (47.8, 72.7) |
| Probability Ranked Best | 0.90% | 0.14% | 20.33% | 78.63% |
| Key: CBD10, cannabidiol 10mg/kg/day; CBD10_CLB, cannabidiol 10mg/kg/day plus clobazam; CBD 20, cannabidiol 20mgmg/kg/day; CBD20_CLB, cannabidiol 20mg/kg/day plus clobazam; FFA0.4, fenfluramine 0.4mg/kg/day up to a max. 17mg/day; FFA0.7, fenfluramine 0.7mg/kg/day up to a max. 26mg/day; MCSF, monthly convulsive seizure frequency. Probability Rank 1, Bayesian probability of regimen in column being ranked the most effective therapy amongst the therapies in the trial network. |  |  |  |  |

**Table 2.**
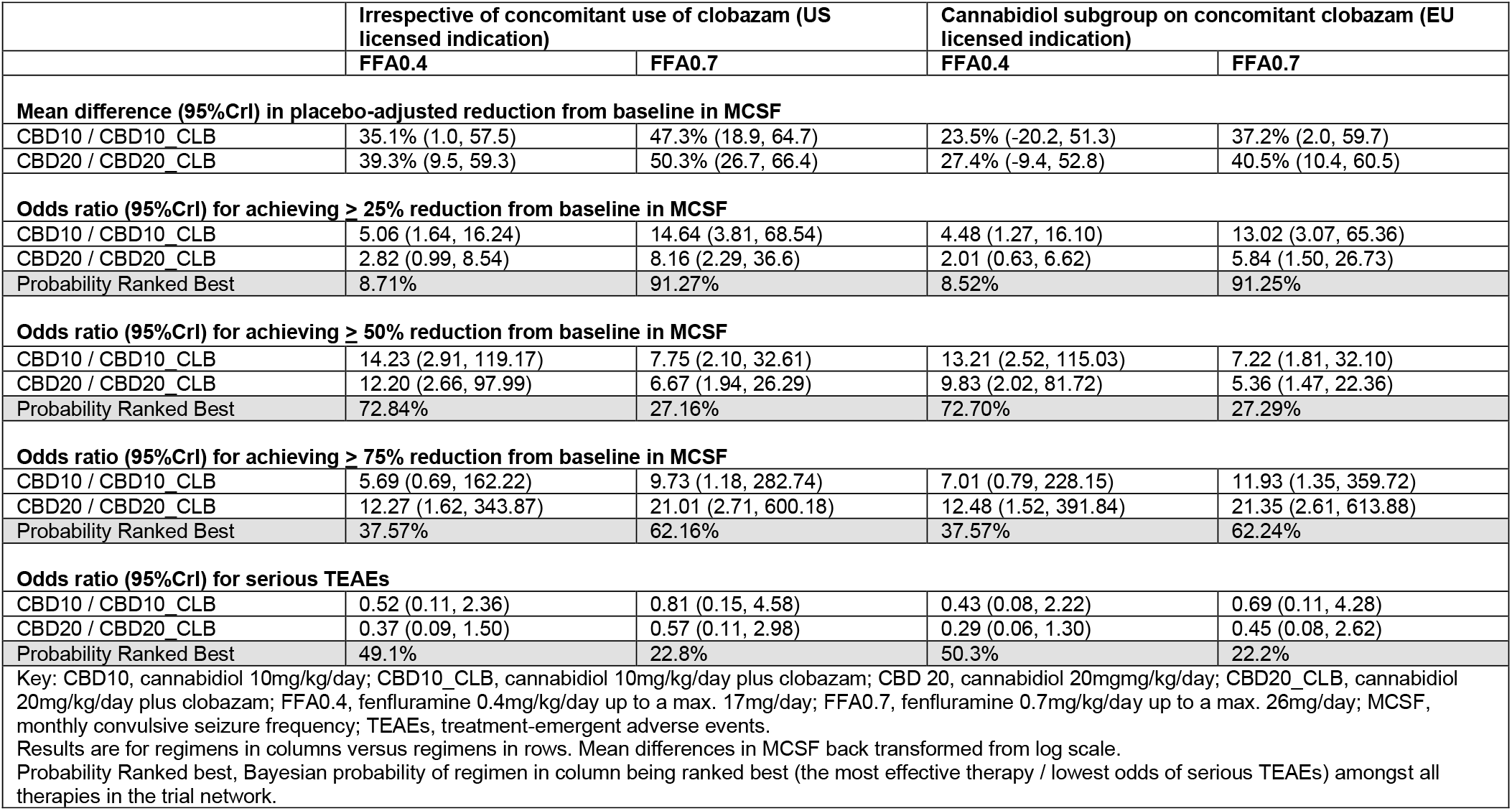
Bayesian Indirect treatment comparisons of fenfluramine versus cannabidiol licensed dose regimens.

When comparing fenfluramine 0.7mg/kg/day and 0.4mg/kg/day versus cannabidiol 10mg/kg/day plus clobazam, the mean differences in placebo-adjusted reduction from baseline in MCSF were 37.2% (2.0, 59.7) and 23.5% (−20.2, 51.3), respectively, in favour of fenfluramine. The mean differences versus cannabidiol 20mg/kg/day plus clobazam were 40.5% (10.4, 60.5) and 27.4% (−9.4, 52.8), respectively, in favour of fenfluramine (**Table 2**). In all comparisons of mean reductions from baseline in MCSF, Bayesian treatment ranking indicated ≥99% probability that fenfluramine regimens were the most effective therapies in the trial networks (**Supplementary Information Figure S3**).

Bucher pairwise ITCs are consistent with these results, demonstrating statistically significant (p≤0.005) additional 33-40% reductions in MCSF for fenfluramine 0.7mg/kg/day compared with cannabidiol 10 and 20mg/kg/day irrespective of clobazam use, and statistically significant (p<0.05) additional 25-32% reductions in MCSF for fenfluramine 0.7mg/kg/day compared with cannabidiol 10 and 20mg/kg/day in the subgroup taking cannabidiol plus clobazam in study GWPCARE2. Fenfluramine 0.4mg/kg/day was statistically (p<0.05) or numerically superior in all comparisons against cannabidiol (**Supplementary Information Table S8**).

### Responder rates: ≥25%, ≥50%, ≥75% and 100% reductions in monthly convulsive seizure frequency

The odds ratios versus placebo for achieving ≥25%, ≥50%, ≥75% reductions from baseline in MCSF showed a clear trend in favour of fenfluramine regimens, both when cannabidiol was used irrespective of concomitant clobazam and when cannabidiol was used with concomitant clobazam (**Figure 2**).

**Figure 2.**
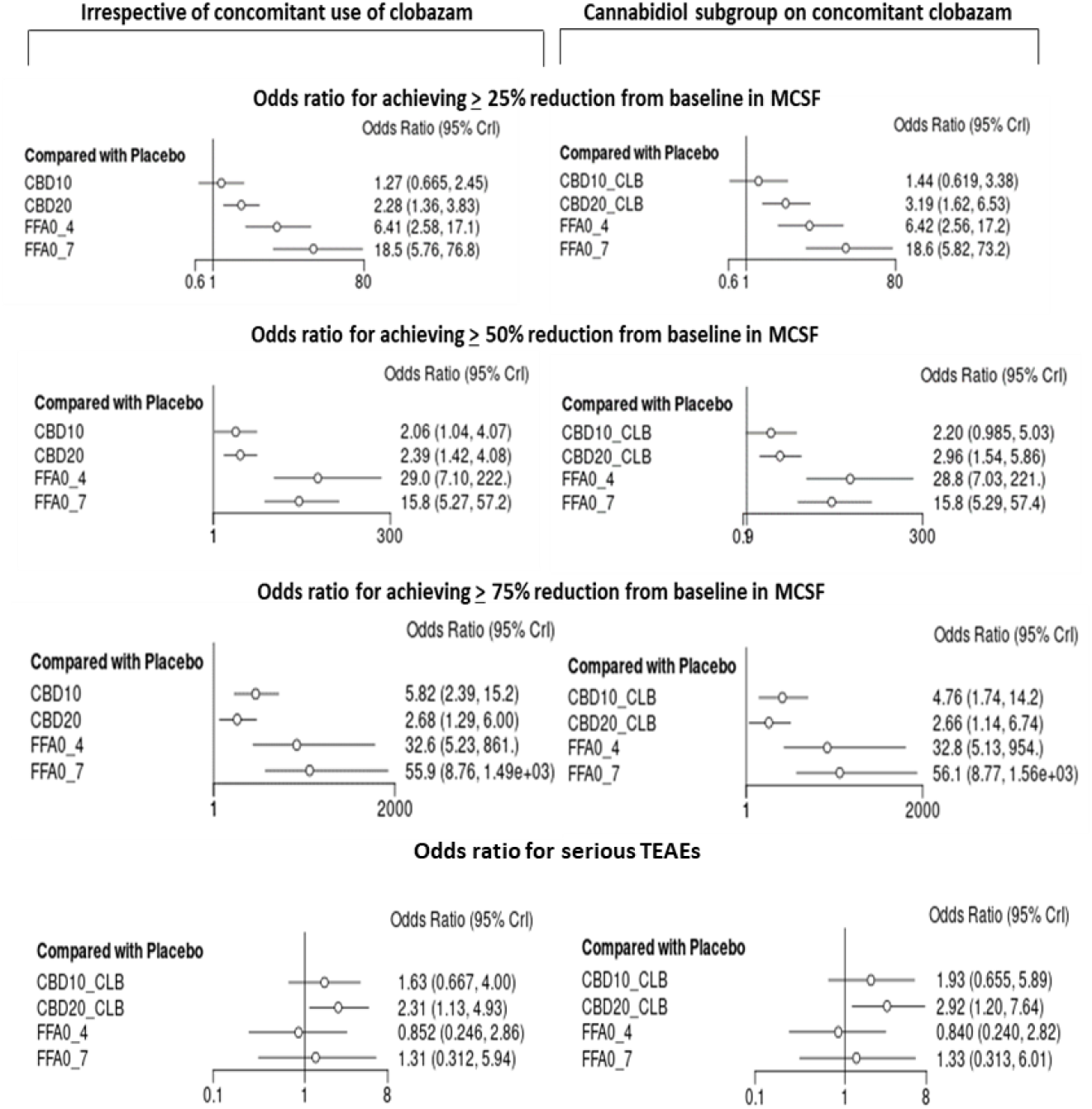
Bayesian NMA of 25%, 50% and 75% responder rates and serious TEAEs. Key: CBD10, cannabidiol 10mg/kg/day; CBD10_CLB, cannabidiol 10mg/kg/day plus clobazam; CBD20, cannabidiol 20mg/kg/day; CBD20_CLB, cannabidiol 20mg/kg/day plus clobazam; FFA_04, fenfluramine 0.4mg/kg/day up to a max. 17mg/day; FFA_07, fenfluramine 0.7mg/kg/day up to a max. 26mg/day

In NMAs indirectly comparing fenfluramine versus cannabidiol, the odds of achieving a clinically meaningful (≥50%) reduction from baseline in MCSF with fenfluramine were 5 to 14 times greater than those with cannabidiol across all licensed dose regimens, and were 5 to 21 times greater for achieving a profound (≥75%) reduction. In all of these comparisons, Bayesian treatment ranking probabilities indicated ≥99% probability that fenfluramine regimens were the most effective therapies in the trial network (**Table 2**). In the Bucher pairwise ITCs, statistically significant (p<0.05) odds ratios for clinically meaningful (≥50%) reductions from baseline in MCSF for fenfluramine versus cannabidiol were observed across all licensed dose regimens (**Supplementary Information Table S8**).

As few patients achieved convulsive seizure freedom during the RCTs (maximum of 7.5% with fenfluramine 0.7mg/kg/day in Study 1, and 0% on placebo in three of the four trials; see **Supplementary Information Figure S4**), formal indirect comparisons were not conducted for 100% responder rates.

### Adverse events

A similar majority of trial participants experienced a TEAE of any severity with fenfluramine and cannabidiol. The incidence of decreased appetite was greater with fenfluramine than cannabidiol (38-44% versus 17-34%) but the reported incidence of decreased weight was similar to cannabidiol (5%). Somnolence or lethargy was reported more frequently with cannabidiol, particularly in those taking concomitant clobazam (36-45%), than with fenfluramine (14-28%). There were no cases of valvular heart disease or pulmonary arterial hypertension with fenfluramine (not reported for cannabidiol). Across cannabidiol regimens 4-11% experienced raised liver enzymes with 4-5% on cannabidiol 20mg/kg/day experiencing abnormal liver function (not reported for fenfluramine) (see **Supplementary information Table S9**).

The incidence of any serious TEAEs was numerically lower with fenfluramine than cannabidiol (**Figure 2**), with reduced odds for fenfluramine versus cannabidiol across all licensed regimens (**Table 2**). Bayesian treatment ranking probabilities indicated a 72% probability of fenfluramine regimens being ranked best (lowest odds of serious TEAEs), and a 71-80% probability of cannabidiol regimens being ranked worst (highest odds of serious TEAEs) among therapies in the trial network (**Supplementary Information Figure S5**). As odds ratios of serious TEAEs were not reported in publications, Bucher pairwise ITCs were not conducted for this outcome.

### Sensitivity analyses

Frequentist NMAs and random effects models generated treatment effect estimates similar to those estimated with the Bayesian fixed effects NMAs. Tests of the available direct and indirect evidence in the trial network indicate no significant inconsistencies. Due to limited reporting in the poster, it was not possible to include fenfluramine Study 3 in the analysis of the placebo-adjusted mean reduction in MCSF; however, the point estimate for this outcome (64.8%) was highly consistent with that reported in Study 1 (62.3%), and a higher proportion (13%) achieved seizure freedom than in any of the other fenfluramine or cannabidiol studies. Inclusion of Study 3 increased the odds ratio for a clinically meaningful (≥50%) reduction from baseline in MCSF with fenfluramine (**Supplementary Table S10)**. Limiting the analyses to fully published data may, therefore, be conservative for fenfluramine.

## Discussion

To our knowledge, this is the first study to assess the comparative efficacy and safety of fenfluramine against cannabidiol across their full US and European licensed dose regimens as add-on therapies in DS. A recent NMA by Devi et al 2021 indirectly compared fenfluramine (0.7mg/kg/day and 0.4mg/kg/day combined) against cannabidiol 20mg/kg/day irrespective of concomitant clobazam use, and concluded that fenfluramine was more effective with a lower incidence of serious TEAEs (30). Our study concurs with these findings and further indicates that fenfluramine in each of its licensed regimens provides superior convulsive seizure control and is well tolerated compared with all licensed dose regimens of cannabidiol, including when cannabidiol is specifically used with concomitant clobazam.

Across multiple convulsive seizure outcomes, including reductions in mean MCSF and clinically meaningful (≥50%) and profound (≥75%) reductions in MCSF, fenfluramine was consistently ranked superior to cannabidiol licensed dose regimens. Given that add-on cannabidiol itself offers a significant improvement in convulsive seizure control compared with continued SoC AEDs in people with DS with treatment-resistant seizures (16,17), these consistent findings of a clear additional benefit in seizure control with fenfluramine are particularly noteworthy.

Qualitative assessment of key adverse events of interest reported in the published RCTs suggests that fenfluramine may be associated with lower rates of somnolence or lethargy and a higher incidence of decreased appetite. However, there was no evidence of an increase in weight loss with fenfluramine compared with that recorded in the cannabidiol trial publications, and with long term treatment fenfluramine has no additional impact on weight or growth compared with that observed generally in people with DS(34). As the licensed indications for both fenfluramine and cannabidiol include a similar need for monitoring weight or growth (11–14), decreased appetite and weight loss do not appear to be significant differentiators for these add-on therapies.

Fenfluramine carries an additional warning of a risk of valvular heart disease and pulmonary hypertension, and requires cardiac monitoring prior to starting, during, and upon discontinuation of treatment (13,14). This is due to a historical association of fenfluramine with these adverse events when used at far higher doses in the treatment of obesity in adults. Importantly, at the licensed doses in patients with DS, no cases of valvular heart disease or pulmonary hypertension have been observed in RCTs (19,20,32), nor with up to 3 years of follow up in an open label extension study (35), nor in longer term prospective (36) and retrospective (37,38)real-world evidence studies. In contrast, the increased risk of hepatocellular injury noted with cannabidiol, particularly when used with concomitant valproate, is based on contemporary data in the DS population (11,12,16,17).

Based on quantitative assessment, the odds of experiencing serious TEAEs (typically defined as medically significant, or life threatening or resulting in death, or requiring hospitalisation or prolongation of hospitalisation) in the RCTs was consistently numerically lower with fenfluramine licensed regimens than with cannabidiol licensed regimens. Fenfluramine is therefore comparatively well tolerated.

### Strengths and limitations

Robust indirect comparisons of fenfluramine and cannabidiol were conducted using Bayesian NMAs of all relevant data from high quality, similarly designed RCTs identified using systematic review methods. Where data allowed, these NMAs were supported by Bucher pairwise ITCs. The convulsive seizure outcomes explored in the indirect comparisons are aligned with the key outcomes used to determine the efficacy of antiseizure agents in both regulatory and clinical practice settings. The results are therefore reliable and informative for people with DS, their carers, and clinicians and other decision-makers.

There are, however, some caveats and limitations. As DS is a rare disease these analyses are based on relatively small RCTs. This can lead to wide credible/confidence intervals around point estimates of treatment effects. Nonetheless, these data consistently indicate fenfluramine on average provides superior seizure control versus cannabidiol across all licensed regimens.

The fenfluramine and cannabidiol RCTs involved 14-15 weeks of comparative treatment, which may be considered relatively short in the context of this life-long syndrome. Both fenfluramine and cannabidiol have longer-term data available from open-label extension (OLE) studies (Study 1503 (14,35)and GWPCARE5 (12,39), respectively). It is not possible to conduct formal comparisons of these longer-term data due to a lack of a common control arm in these OLE studies. However, as the fenfluramine OLE study indicates its treatment effects are maintained, with no evidence of a waning of effect over long-term treatment up to 3 years (35), the superior efficacy of fenfluramine over cannabidiol demonstrated in the indirect comparisons of RCT data may plausibly be maintained over the long term.

The indirect comparisons of efficacy were focused on convulsive seizure outcomes only. Although non-convulsive seizures are also important outcomes for people with DS and their carers, they are more difficult to detect and report accurately in RCTs, and so would provide a less robust outcome measure for indirect comparison. Status epilepticus and SUDEP events, as the primary causes of premature death in DS (5,6), are of paramount importance; however, RCTs in DS could never be powered adequately for these outcomes. In a recently published pooled analysis of 732 people with DS treated with fenfluramine across RCTs, open-label extension studies and early access programs, the observed incidence rates of all-cause and SUDEP mortality (both 1.7 per 1000 patient years) were substantially lower than those reported in the literature with SoC therapy (15.8 and 9.3 per 1000 person-years, respectively), which the authors attributed largely to the substantial additional reductions in convulsive seizure frequency observed with fenfluramine (24). But in the absence of comparative data it is not possible to estimate an indirect treatment effect on these outcomes in the current study.

NMAs rely on the assumption there is no heterogeneity in effect modifiers across the trials included in the network (40). A detailed NMA feasibility assessment indicated that the cannabidiol and fenfluramine trial designs and participants were on average sufficiently similar to permit NMAs; however, given the heterogenous DS populations within each of the trials, and the need to tailor treatment to individual needs (10), either drug may prove to be more effective in an individual patient. There are potentially important differences in concomitant AEDs received by participants in the RCTs. Clobazam is a significant effect modifier of cannabidiol, as reflected in the cannabidiol label in Europe that mandates its concomitant use(12). For this reason, NMAs were conducted in both the overall trial populations (irrespective of concomitant clobazam use) and using subgroup data from patients taking cannabidiol with concomitant clobazam. As clobazam is not a significant effect modifier of fenfluramine (15) it is appropriate to use the whole fenfluramine trial population data in both comparisons with cannabidiol.

Stiripentol has a pharmacokinetic interaction with fenfluramine, and for this reason fenfluramine is dosed at up to 0.7mg/kg/day (maximum 26mg/day) without concomitant stiripentol (as in Study 1 and Study 3), and at up to 0.4mg/kg/day (maximum 17mg/day) with concomitant stiripentol (as in Study 2) (13,14). Of note, stiripentol and clobazam exposure is increased by cannabidiol (12) and significant proportions of participants in the cannabidiol trials were receiving concomitant stiripentol (up to 58% in those also receiving clobazam) (18). However, as there are no publicly available efficacy and safety data for cannabidiol based on concomitant stiripentol use, it was not possible to conduct separate NMAs for fenfluramine versus cannabidiol based on stiripentol use. Both licensed doses of fenfluramine were therefore pragmatically included separately in the same networks for comparisons against cannabidiol. As Bucher pairwise ITCs confirmed the results of the NMAs, this pragmatic approach appears to be justified.

Finally, stiripentol is itself licensed as an add-on to standard of care AEDs in DS (41,42) but was excluded as a comparator from our analyses. Stiripentol has been available in Europe for many years and is regarded as a SoC AED in DS (43). Furthermore, there was a high proportion of concomitant use of stiripentol in the cannabidiol RCTs (18) and fenfluramine Study 2 provides efficacy data for fenfluramine when added on to stiripentol (20), which precludes their use in a comparative analysis. Fenfluramine Study 1 provides efficacy data in stiripentol naïve patients (52.3%) and stiripentol experienced patients no longer taking stiripentol (48.7%), and post hoc analysis indicates that fenfluramine efficacy is highly consistent in both these subgroups (19,44). However, there are other challenges in conducting robust indirect comparisons with these subgroup data. A Cochrane systematic review (45) noted that stiripentol is supported by two small RCTs (STICLO-France [n=41] and STICLO-Italy [n=23]) (46,47), which were of only 8 weeks duration, assessed outcomes in only the last 4 weeks of treatment and were deemed to provide only low to moderate quality evidence (45). Collectively, our exclusion of stiripentol is therefore appropriate. Nonetheless, despite these significant issues, the recent NMA by Devi et al 2021 included stiripentol and concluded fenfluramine had comparable efficacy but with less frequent serious TEAEs (30).

## Conclusion and implications

Fenfluramine provides superior convulsive seizure control and is comparatively well tolerated versus cannabidiol in all its licensed dose regimens when added to SoC AEDs in DS. Fenfluramine may meet the need for a highly effective and tolerable add-on therapy to control seizures in Dravet syndrome.

## Data Availability

Zogenix International Ltd does not currently have a data sharing policy. All data used in the analyses are available in the cited references.

