## Supplementary materials for "Comparative effectiveness of fenfluramine versus cannabidiol in their licensed indications for the treatment of seizures in Dravet Syndrome: a systematic review and network meta-analysis"

*Table S 1. Sample of search strategy (PubMed)*

| **Search strategy** | | **Hits (to 28 Jun 2020)** | **Hits (to 30 Nov 2021)** |
|---|---|---|---|
| #1 | "Epilepsies, Myoclonic"[Mesh] | 4627 | 5064 |
| #2 | (child OR childhood OR children OR infan*) | 3225849 | 3480975 |
| #3 | #1 AND #2 | 2256 | 2477 |
| #4 | "dravet syndrome" | 940 | 1209 |
| #5 | "childhood epileptic encephalopathy" | 30 | 33 |
| #6 | "severe myoclonic epilepsy" | 368 | 378 |
| #7 | SMEI | 171 | 180 |
| #8 | Dravet* | 1254 | 1537 |
| #9 | "dravet's syndrome" | 38 | 40 |
| #10 | "childhood epileptic encephalopathies" | 17 | 22 |
| #11 | "childhood epilepsy encephalopathies" | 6506 | 7065 |
| #12 | "childhood epilepsy encephalopathy" | 6506 | 7065 |
| #13 | #3 OR #4 OR #5 OR #6 OR #7 OR #8 OR #9 OR #10 OR #11 OR #12 | 9067 | 9964 |
| #14 | randomized controlled trial [pt] | 509383 | 553463 |
| #15 | controlled clinical trial [pt] | 596182 | 643146 |
| #16 | randomized [tiab] | 523366 | 591345 |
| #17 | placebo [tiab] | 214300 | 230241 |
| #18 | clinical trials as topic [mesh: noexp] | 341863 | 367152 |
| #19 | randomly [tiab] | 335824 | 372584 |
| #20 | trial [ti] | 219812 | 252685 |
| #21 | #14 OR #15 OR #16 OR #17 OR #18 OR #19 OR #20 | 1378631 | 1504339 |
| #22 | #13 AND #21 | 428 | 505 |
| #23 | animals [mh] NOT humans [mh] | 4712175 | 4923607 |
| #24 | #22 NOT #23 | 427 | 504 |
| #25 | English[lang] | 26448823 | 28663084 |
| #26 | #24 AND #25 | **396** | 472 |

| Search strategy | | Hits (to 28 Jun 2020) | Hits (to 30 Nov 2021) |
|---|---|---|---|
| #27 | #24 AND #25 from 2020/6/29 - 2021/11/30 | **-** | **66** |

*Table S 2. Conference proceedings included in searches*

| **Conference meetings (2017-2021)** |
|---|
| American Epilepsy Society (AES) annual meetings |
| British Paediatric Neurology Association meetings |
| European Congress on Epileptology meetings |
| European Paediatric Neurology Society Congress meetings |
| International Epilepsy Congress meetings |

*Table S 3. Eligibility criteria applied in screening*

| | **Inclusion criteria** |
|---|---|
| Population | • All patients with a defined clinical diagnosis of DS (with or without confirmed SCN1A mutation), irrespective of age.<br>• If studies include mixed populations of patients, only those reporting results separately for DS patients will be included |
| Intervention(s) | • Fenfluramine (synonym: ZX008) licensed doses of 0.7 mg/kg/day, or 0.4 mg/kg/day if co-administered with stiripentol, given as an add-on therapy to standard of care AEDs. This will include fenfluramine as the hydrochloride salt at doses of 0.8mg/kg/day, or 0.5 mg/kg/day if co-administered with stiripentol.<br>• Cannabidiol (synonym: GWP42003-P) in the form of the highly purified cannabidiol used in the Epidyolex/Epidiolex formulation, at licensed doses of up to 10mg/kg/day or 20mg/kg/day |
| Comparator(s) | • Any active pharmacological comparator used as add-on therapy to standard of care AEDs<br>• Placebo/standard of care AEDs |
| Outcomes | • Any outcome measure that aligns to the following measurements of clinical effectiveness or adverse events/tolerability of AEDs in the management of seizures in DS:<br> • Change in convulsive seizure frequency from baseline<br> • Responder rate (≥25%, ≥50% or ≥75% reduction from baseline in convulsive seizure frequency)<br> • Serious adverse events rates, individual adverse events & tolerability |
| Study design | • Open-label or blinded RCTs involving at least 10 subjects per treatment arm – including re-analyses of RCTs<br>• Systematic literature reviews – for background information, reference checking and RCT data not provided in primary publications |

| | |
|---|---|
| | • HTA reports providing RCT data not included in primary publications |
| Geographic coverage | • Any geographic location |

*Table S 4. PRISMA NMA Checklist of Items to Include When Reporting A Systematic Review Involving a Network Meta-analysis*

| Section/Topic | Item # | Checklist Item | Reported on Page # |
|---|---|---|---|
| **TITLE** | | | |
| Title | 1 | Identify the report as a systematic review *incorporating a network meta-analysis (or related form of meta-analysis).* | p1 |
| **ABSTRACT** | | | |
| Structured summary | 2 | Provide a structured summary including, as applicable:<br>**Background:** main objectives<br>**Methods:** data sources; study eligibility criteria, participants, and interventions; study appraisal; and *synthesis methods, such as network meta-analysis.*<br>**Results:** number of studies and participants identified; summary estimates with corresponding confidence/credible intervals; *treatment rankings may also be discussed. Authors may choose to summarize pairwise comparisons against a chosen treatment included in their analyses for brevity.*<br>**Discussion/Conclusions:** limitations; conclusions and implications of findings.<br>**Other:** primary source of funding; systematic review registration number with registry name. | p1 |
| **INTRODUCTION** | | | |
| Rationale | 3 | Describe the rationale for the review in the context of what is already known*, including mention of why a network meta-analysis has been conducted.* | p4 |
| Objectives | 4 | Provide an explicit statement of questions being addressed, with reference to participants, interventions, comparisons, outcomes, and study design (PICOS). | p3-4 |
| **METHODS** | | | |
| Protocol and registration | 5 | Indicate whether a review protocol exists and if and where it can be accessed (e.g., Web address); and, if available, provide registration information, including registration number. | p4 |
| Eligibility criteria | 6 | Specify study characteristics (e.g., PICOS, length of follow-up) and report characteristics (e.g., years considered, language, publication status) used as criteria for eligibility, giving rationale. *Clearly describe eligible treatments included in the treatment network, and note whether any have been clustered or merged into the same node (with justification).* | Table S3 |
| Information sources | 7 | Describe all information sources (e.g., databases with dates of coverage, contact with study authors to identify additional studies) in the search and date last searched. | p4 |

| | | | |
|---|---|---|---|
| Search | 8 | Present full electronic search strategy for at least one database, including any limits used, such that it could be repeated. | Table S1 |
| Study selection | 9 | State the process for selecting studies (i.e., screening, eligibility, included in systematic review, and, if applicable, included in the meta-analysis). | p4, Table S3 |
| Data collection process | 10 | Describe method of data extraction from reports (e.g., piloted forms, independently, in duplicate) and any processes for obtaining and confirming data from investigators. | p4 |
| Data items | 11 | List and define all variables for which data were sought (e.g., PICOS, funding sources) and any assumptions and simplifications made. | Table S5<br>Table S6<br>Table S7 |
| **Geometry of the network** | **S1** | Describe methods used to explore the geometry of the treatment network under study and potential biases related to it. This should include how the evidence base has been graphically summarized for presentation, and what characteristics were compiled and used to describe the evidence base to readers. | Fig 1 |
| Risk of bias within individual studies | 12 | Describe methods used for assessing risk of bias of individual studies (including specification of whether this was done at the study or outcome level), and how this information is to be used in any data synthesis. | p4, Fig S2 |
| Summary measures | 13 | State the principal summary measures (e.g., risk ratio, difference in means). *Also describe the use of additional summary measures assessed, such as treatment rankings and surface under the cumulative ranking curve (SUCRA) values, as well as modified approaches used to present summary findings from meta-analyses.* | p5 |
| Planned methods of analysis | 14 | Describe the methods of handling data and combining results of studies for each network meta-analysis. This should include, but not be limited to:<br>• *Handling of multi-arm trials;*<br>• *Selection of variance structure;*<br>• *Selection of prior distributions in Bayesian analyses; and*<br>• *Assessment of model fit.* | Meta-insight, p5 |
| **Assessment of Inconsistency** | **S2** | Describe the statistical methods used to evaluate the agreement of direct and indirect evidence in the treatment network(s) studied. Describe efforts taken to address its presence when found. | p5, Table S10 |
| Risk of bias across studies | 15 | Specify any assessment of risk of bias that may affect the cumulative evidence (e.g., publication bias, selective reporting within studies). | p5-6 |
| Additional analyses | 16 | Describe methods of additional analyses if done, indicating which were pre-specified. This may include, but not be limited to, the following:<br>• Sensitivity or subgroup analyses;<br>• Meta-regression analyses;<br>• *Alternative formulations of the treatment network; and*<br>• *Use of alternative prior distributions for Bayesian analyses (if applicable).* | p5, p8 |
| **RESULTS†** | | | |
| Study selection | 17 | Give numbers of studies screened, assessed for eligibility, and included in the review, with reasons for exclusions at each stage, ideally with a flow diagram. | p6, Fig S1 |
| **Presentation of network structure** | **S3** | Provide a network graph of the included studies to enable visualization of the geometry of the treatment network. | Fig 1 |
| **Summary of network geometry** | **S4** | Provide a brief overview of characteristics of the treatment network. This may include commentary on the abundance of trials and randomized patients for the different interventions and pairwise comparisons in the network, gaps of evidence in the treatment network, and potential biases reflected by the network structure. | p6 |

| | | | |
|---|---|---|---|
| Study characteristics | 18 | For each study, present characteristics for which data were extracted (e.g., study size, PICOS, follow-up period) and provide the citations. | Table S5<br>Table S6<br>Table S7 |
| Risk of bias within studies | 19 | Present data on risk of bias of each study and, if available, any outcome level assessment. | p5, Fig S2 |
| Results of individual studies | 20 | For all outcomes considered (benefits or harms), present, for each study: 1) simple summary data for each intervention group, and 2) effect estimates and confidence intervals. *Modified approaches may be needed to deal with information from larger networks.* | Table S7<br>Table S9 |
| Synthesis of results | 21 | Present results of each meta-analysis done, including confidence/credible intervals. *In larger networks, authors may focus on comparisons versus a particular comparator (e.g. placebo or standard care), with full findings presented in an appendix. League tables and forest plots may be considered to summarize pairwise comparisons.* If additional summary measures were explored (such as treatment rankings), these should also be presented. | Table 1<br>Table 2<br>Fig 2<br>Fig S3<br>Fig S5 |
| **Exploration for inconsistency** | **S5** | Describe results from investigations of inconsistency. This may include such information as measures of model fit to compare consistency and inconsistency models, *P* values from statistical tests, or summary of inconsistency estimates from different parts of the treatment network. | p8,<br>Table S10 |
| Risk of bias across studies | 22 | Present results of any assessment of risk of bias across studies for the evidence base being studied. | p5, Fig S2 |
| Results of additional analyses | 23 | Give results of additional analyses, if done (e.g., sensitivity or subgroup analyses, meta-regression analyses*, alternative network geometries studied, alternative choice of prior distributions for Bayesian analyses,* and so forth). | p8,<br>Table S10 |
| **DISCUSSION** | | | |
| Summary of evidence | 24 | Summarize the main findings, including the strength of evidence for each main outcome; consider their relevance to key groups (e.g., healthcare providers, users, and policy-makers). | p8-12 |
| Limitations | 25 | Discuss limitations at study and outcome level (e.g., risk of bias), and at review level (e.g., incomplete retrieval of identified research, reporting bias). *Comment on the validity of the assumptions, such as transitivity and consistency. Comment on any concerns regarding network geometry (e.g., avoidance of certain comparisons).* | p9-12 |
| Conclusions | 26 | Provide a general interpretation of the results in the context of other evidence, and implications for future research. | p12 |
| **FUNDING** | | | p12 |
| Funding | 27 | Describe sources of funding for the systematic review and other support (e.g., supply of data); role of funders for the systematic review. This should also include information regarding whether funding has been received from manufacturers of treatments in the network and/or whether some of the authors are content experts with professional conflicts of interest that could affect use of treatments in the network. | |

PICOS = population, intervention, comparators, outcomes, study design.
* Text in italics indicates wording specific to reporting of network meta-analyses that has been added to guidance from the PRISMA statement.
† Authors may wish to plan for use of appendices to present all relevant information in full detail for items in this section.

*Table S 5. Comparability of study designs, eligibility criteria and endpoint assessment*

| | **Study** | | | | |
|---|---|---|---|---|---|
| | **Fenfluramine Study 1** (19) | **Fenfluramine Study 2** (20) | **Fenfluramine Study 3** (32) | **Cannabidiol GWPCARE1 Part B** (16) | **Cannabidiol GWPCARE2** (17) |
| **Intervention and comparator** | FFA 0.7mg/kg/day (max. 26mg/day) or FFA 0.2mg/kg/day or placebo | FFA 0.4mg/kg/day (max. 17mg/day) or placebo | FFA 0.7mg/kg/day (max. 26mg/day) or FFA 0.2mg/kg/day or placebo | CBD 20mg/kg/day or placebo | CBD 10mg/kg/day or CBD 20mg/kg/day or placebo |
| **Study design and size** | Phase 3 placebo-controlled RCT (n=119) | Phase 3 placebo-controlled RCT (n=87) | Phase 3 placebo-controlled RCT (n=143) | Phase 3 placebo-controlled RCT (n=120 in whole trial; n=78 in the licensed subpopulation taking CLB) | Phase 3 placebo-controlled RCT (n=198 in the whole trial; n=126 in the licensed subpopulation taking CLB) |
| **Year(s) of conduct** | 2016–2018 | 2016–2018 | 2016-2020 | 2015 | 2015–2018 |
| **Study and treatment duration** | 6-week baseline, **2-week titration + 12-week maintenance** | 6-week baseline, **3-week titration + 12-week maintenance** | 6-week baseline, **2- week titration + 12- week maintenance** | 4-week baseline, **2- week titration + 12- week maintenance** | |
| **Eligibility** | • DS<br>• 2–18 years old<br>• ≥4 convulsive seizures per 4- week period during previous 12 weeks prior to screening<br>≥6 convulsive seizures during 42-day baseline with ≥2 in first 3 weeks and ≥2 in last 3 weeks | | • DS<br>• 2–18 years old | • DS<br>• 2–18 years old<br>≥4 convulsive seizures during 28-day baseline | |

| | **Study** | | | | |
|---|---|---|---|---|---|
| | **Fenfluramine Study 1** (19) | **Fenfluramine Study 2** (20) | **Fenfluramine Study 3** (32) | **Cannabidiol GWPCARE1 Part B** (16) | **Cannabidiol GWPCARE2** (17) |
| **Background medication** | • No STP in the 21 days prior to screening<br>• One or more stable AEDs<br>• All other medications or interventions must be stable for ≥4 weeks prior to screening and are expected to remain stable throughout the study | • Receiving stable dose of CLB, and/or VPA, and 100% STP<br>• All medications or interventions for epilepsy must be stable for ≥4 weeks prior to screening and are expected to remain stable throughout the study | • No STP in the 21 days prior to screening<br>• One or more stable AEDs | • One or more stable AEDs<br>• All medications or interventions must be stable for ≥4 weeks prior to screening and are expected to remain stable throughout the study | |
| **Endpoints** | | | | | |
| **Convulsive seizure definition** | Generalised tonic-clonic, tonic, clonic, tonic-atonic, hemiclonic, and focal seizures with an observable motor component | | NS | Tonic, clonic, tonic-clonic or atonic | |
| **Reduction in convulsive seizures** | % change in CSF between baseline and T+M periods (per 28 days) (primary endpoint)<br>Parametric assessment of % reduction from baseline in convulsive seizure frequency per 28 days compared with placebo (i.e. the additional reduction over placebo) | | | % change in CSF between baseline and T+M periods (per 28 days) (primary endpoint)<br>Parametric assessment of % reduction from baseline in convulsive seizure frequency per 28 days compared with placebo (i.e. the additional reduction over placebo) | |
| **Responder rates (proportion achieving at least 25, 50 or 75% reduction from baseline in convulsive seizure frequency** | Reported on CSF over combined T+M period (per 28 days) vs baseline (key secondary endpoint) | | | Reported on CSF over combined T+M period (per 28 days) (key secondary endpoint) | |
| **Serious TEAEs** | Reported over combined T+M period | | | Reported over combined T+M period | |
| **Key:** AED, antiepileptic drug; CBD, cannabidiol; CLB, clobazam; CSF, convulsive seizure frequency; FFA, fenfluramine; NR, not reported; RCT, randomized controlled trial; STP, stiripentol; T+M, titration and maintenance treatment period; VPA, valproate | | | | | |

*Table S 6. Comparison of baseline characteristics across RCTs*

| | RCT | | | | | | |
|---|---|---|---|---|---|---|---|
| | **Fenfluramine** | | | **Cannabidiol** | | | |
| | **Study 1** (19) | **Study 2 (previously 1504)** (20) | **Study 3** (32) | **Cannabidiol GWPCARE1B Full population** (16) | **Cannabidiol GWPCARE2 Full population** (17) | **Cannabidiol 10mg/kg/day Clobazam subgroup** (18) | **Cannabidiol 20mg/kg/day Clobazam subgroup** (18) |
| **N** | 119 | 87 | 143 | 120 (full population) | 198 (full population) | 45 | 80 |
| **Age – year (mean±SD)** | 9.0 ±4.65 | 9.1 ±4.80 | ~9 | 9.8 ±4.8 | 9.3 ±4.4 | 9.1±4.1 | 9.3±4.3 |
| **Sex – no. (% male)** | 64 (54) | 50 (57.5) | 74 (51.7) | 62 (52) | 94 (47.5) | 23 (51) | 45 (56) |
| **BMI – kg/m$^2$ (mean±SD)** | 18.57 ±4.408 | 18.24 ±4.049 | NR | 18.7±4.6 | 18.7±4.3 | 18.9 | 19.0 |
| **White** | 98 (82.4) | 52 (59.8) | 106 (74.1) | 94 (78.3) | 176 (88.9) | NR | NR |
| **Black or African American** | NR | 3 (3.4) | NR | 4 (3.3) | 5 (2.5) | NR | NR |
| **Asian** | 7 (5.9) | 3 (3.4) | 20 (14.0) | 1 (0.8) | 5 (2.5) | NR | NR |
| **American/Alaska Native** | 2 (1.7) | 0 (0) | NR | NR | 1 (0.5) | NR | NR |
| **Other/not reported/unknown** | 12 (10.1) | 29 (33.3) | 16 (11.2) | 21 (17.5) | 11 (5.6) | NR | NR |
| **United States** | 70 (58.8) | 22 (25.3) | NR | 72 (60) | 93 (47) | 22 (49) | 44 (55) |
| **Rest of world** | 49 (41.2) | 65 (74.7) | NR | 48 (40) | 105 (53) | 23 (51) | 36 (45) |
| **No. of previous AEDs** | NR | NR | NR | 4.6±3.8 | Median 4 | Median 4 | Median 4 |
| **No. of concomitant AEDs** | Mean 2.4 | Mean 3.5 | NR | Mean 2.9±1.0 | Median 3 | Median 3 | Median 3 |
| **Convulsive seizures per 28 days (baseline)** | FFA 0.2 mg/kg/day: median 17.5 (range 4.7– 623.5)<br><br>FFA 0.7 mg/kg/day: median 20.7 (range 4.8–124.0)<br><br>Placebo: median 27.3 (range 3.3–147.3) | FFA 0.4 mg/kg/day: median 14.0 (range 2.7– 213.3)<br><br>Placebo: median 10.7 (range 2.7–162.7) | FFA 0.2 mg/kg/day: median 18.0 (range 4.0– 1464)<br><br>FFA 0.7 mg/kg/day: median 13.0 (range 2.7–2701)<br><br>Placebo: median 12.7 (range 4.0–229.3) | CBD 20 mg/kg/day: median 12.4 (range 3.9–1717)<br><br>Placebo: median 14.9 (range 3.7–718) | CBD 20 mg/kg/day: median 9.0 (range 3.9–661.2)<br><br>CBD 10 mg/kg/day: median 13.5 (range 0–467.0)<br><br>Placebo: median 16.6 (range 3.0–770.5) | GWPCARE2: CBD 10 mg/kg/day: median 13.1 (range 4.0–238.4) | GWPCARE1B: CBD 20 mg/kg/day: median 9.6 (range 3.9–661.2)<br><br>GWBCARE2: CBD 20 mg/kg/day: median 10.8 (range 3.9–553,5) |
| **Clobazam** | 70 (58.8) | 82 (94.3) | NR | 78 (65) | 126 (64) | 45 (100) | 80 (100) |
| **Valproate, all forms** | 71 (59.7) | 66 (75.9) | NR | 71 (59) | 139 (70) | 30 (67) | 56 (70) |
| **Stiripentol** | 0 | 87 (100) | NR | 51 (42) | 71 (36) | 17 (38) | 51 (51) |
| **Levetiracetam** | 26 (21.8) | 10 (11.5) | NR | 33 (28) | 54 (27) | 11 (24) | 15 (19) |
| **Topiramate** | 30 (25.2) | 21 (24.1) | NR | 31 (26) | 46 (23) | 8 (18) | 18 (23) |
| **Key:** AED; antiepileptic drug; BMI, body mass index; CBD, cannabidiol; FFA, fenfluramine; NR, not reported; IQR, interquartile range; SD, standard deviation; STP, stiripentol. | | | | | | | |

*Table S 7. Data used in NMAs*

| Trial | Treatment | n | % change from baseline in CSF per 28 days vs placebo | | ≥25% reduction in CSF n/N (%) | ≥50% reduction in CSF n/N (%) | ≥75% reduction in CSF n/N (%) | Serious TEAEs n/N (%) |
|---|---|---|---|---|---|---|---|---|
| | | | **Parametric analysis (95% CI)** | **Log-transformed mean values (95% CI)¶** | | | | |
| **Fenfluramine** | | | | | | | | |
| **Study 1** (19) | Placebo | 40 | Reference | 0.000 | 14/40 (35) | 5/40 (12.5) | 1/40 (2.5) | 4/40 (10.0) |
| | FFA 0.7 mg/kg/day (max 26 mg/day) | 40 | ANCOVA: -62.3 (-72.8, -47.7)* | -0.976 (-1.302, -0.648) | 36/40 (90) | 27/40 (67.5) | 20/40 (50.0) | 5/40 (12.5) |
| **Study 2 (previously Study 1504)** (20) | Placebo | 44 | Reference | 0.000 | 12/44 (27.3) | 2/44 (4.5) | 1/44 (2.3) | 7/44 (15.9) |
| | FFA 0.4 mg/kg/day (max 17mg/day) | 43 | ANCOVA: -54.0 (-67.2, -35.6)* | -0.777 (-1.115, -0.440) | 30/43 (69.8) | 23/43 (53.5) | 15/43 (34.9) | 6/43 (14.0) |
| **Study 3** (32) | Placebo | 48 | Reference | - | 13/48 (27) | 3/48 (6) | 2/48 (4) | 2/48 (4) |
| | FFA 0.7 mg/kg/day (max 26 mg/day) | 49 | ANCOVA*: -64.8* | - | 40/48 (83) | 35/48 (73) | 23/48 (48) | 3/49 (6) |
| **Cannabidiol - full trial populations** | | | | | | | | |
| **GWPCARE1B** (16) | Placebo | 59 | Reference | 0.000 | 26/59 (44.1) | 16/59 (27.1) | 7/59 (11.9) | 3/59 (5.1) |
| | CBD 20 mg/kg/day | 61 | ANCOVA: -21.99 (-45.2, 1.22)□ | -0.248 (-0.601, 0.0012) | 38/61 (62.3) | 26/61 (42.6) | 14/61 (23.0) | 10/61 (16.4) |
| **GWPCARE2** (17) | Placebo | 65 | Reference | 0.000 | 32/65 (49.2) | 17/65 (26.2) | 4/65 (6.2) | 10/65 (15.4) |
| | CBD 10 mg/kg/day | 66 | NBR: -29.8 (-46.2, -8.4) | -0.354 (-0.620, -0.088) | 37/66 (56.1) | 29/66 (43.9) | 20/66 (30.3) | 13/64 (20.3) |
| | CBD 20 mg/kg/day | 67 | NBR: -25.7 (-43.7, -2.9) | -0.297 (-0.574, -0.029) | 47/67 (70.1) | 33/67 (49.3) | 12/67 (17.9) | 17/69 (24.6) |
| **Cannabidiol - subgroup taking clobazam** | | | | | | | | |
| **GWPCARE1B** (18,33) | Placebo | 38 | Reference | 0.000 | 16/38 (42.1) | 9/38 (23.7) | 5/38 (13.2) | 1/38 (2.6) |
| | CBD 20 mg/kg/day | 40 | NBR: -42.8 (-60.4, -17.4)‖ | -0.559 (-0.926, -0.191) | 26/40 (65.0) | 19/40 (47.5) | 10/40 (25.0) | 8/40 (20.0) |
| **GWPCARE2** (18,33) | Placebo | 41 | Reference | 0.000 | 24/41 (58.5) | 15/41 (36.6) | 4/41 (9.8) | 7/41 (17.1) |
| | CBD 10 mg/kg/day | 45 | NBR: -37.4 (-54.5, -13.9)‖ | -0.468 (-0.787, -0.150) | 31/45 (68.9) | 25/45 (55.6) | 16/45 (35.6) | 10/44 (22.7) |
| | CBD 20 mg/kg/day | 40 | NBR: -30.8 (-50.4, -3.6)‖ | -0.368 (-0.701, -0.037) | 34/40 (85.0) | 25/40 (62.5) | 10/40 (25.0) | 11/41 (26.8) |

**Key:** CBD, cannabidiol; CI, confidence interval; CLB, clobazam; CSF, convulsive seizure frequency; FFA, fenfluramine; NBR, negative binomial regression model; TEAEs, treatment-emergent adverse events

¶ Percentage change from parametric analyses converted to relative rates and log transformed as ln(1+ percentage change), along with 95% CIs from which arm-based standard deviations were calculated

* ANCOVA model with treatment group and age group (<6 years, ≥6 years) as factors, baseline CSF as a covariate and % change from baseline CSF during treatment period as response

□Sensitivity analysis based on ANCOVA model, as primary analysis based on median change rather than mean.

‖Data for the clobazam subgroup are based on NBR model that includes total number of seizures as a response variable, age group, time (baseline and treatment period), treatment, and treatment by time interaction as fixed effects, and subject as a random effect. Log-transformed number of days in which seizures were reported by period is included as an offset.

*Table S 8. Bucher ITCs for efficacy outcomes of interest*

| Cannabidiol dataset | Source of cannabidiol data | Cannabidiol vs placebo | Fenfluramine 0.7mg/kg/day (max 26mg/day) vs placebo (19) | Fenfluramine 0.4mg/kg/day (max 17mg/day) vs placebo (20) | ITC: Fenfluramine 0.7mg/kg/day vs Cannabidiol | ITC: Fenfluramine 0.4mg/kg/day vs Cannabidiol |
|---|---|---|---|---|---|---|
| **Mean monthly reduction in CSF vs placebo (% Mean difference, 95%CI) – Full trial population** | | | | | | |
| CBD10 | GWPCARE2 study (17) | 29.8 (8.4, 46.2) | | | **32.5 (9.8, 55.2) p=0.0050** | 24.2 (-0.43, 48.8) p=0.0542 |
| CBD20 | GWPCARE2 study (17) | 25.7 (2.9, 43.2) | 62.3 (47.7, 72.8) | 54.0 (35.6, 67.2) | **36.6 (12.9, 60.3) p=0.0025** | **28.3 (2.7, 53.9) p=0.0303** |
| CBD20 | GWPCARE1 study (16) | 22.0 (-1.2, 45.2)¶ | | | **40.3 (13.9, 66.7) p=0.0027** | **32.0 (3.9, 60.1) p=0.0255** |
| **Mean monthly reduction in CSF vs placebo (% Mean difference, 95%CI) – Cannabidiol subgroup taking clobazam** | | | | | | |
| CBD10 _CLB | | 37.4 (13.9, 54.5) | | | **24.9 (1.03, 48.8) p=0.0409** | 16.6 (-9.1, 42.3) p=0.2059 |
| CBD20_CLB (GWPCARE2) | Gunning et al 2020 (18) | 30.8 (3.6, 50.4) | 62.3 (47.7, 72.8) | 54.0 (35.6, 67.2) | **31.5 (5.0, 58.1) p=0.0201** | 23.2 (-5.0, 51.4) p=0.1073 |
| CBD20_CLB (GWPCARE1B) | | 42.8 (17.4, 60.4) | | | 19.5 (-5.4, 44.4) p=0.1247 | 11.2 (-15.5, 37.9) p=0.4107 |
| **≥25% Responder rates (Odds ratios, 95%CI) – Full trial population*** | | | | | | |
| CBD10 | GWPCARE2 study (17) | 1.3 (0.7, 2.6) | | | **17.2 (3.9, 74.9) p=0.0002** | **4.92 (1.6, 15.5) p=0.0066** |
| CBD20 | GWPCARE2 study (17) | 2.4 (1.2, 5.0) | 22.3 (6.0, 84.0) | 6.4 (2.5, 16.5) | **9.3 (2.1, 41.7) p=0.0036** | 2.67 (0.8, 8.7) p=0.1041 |
| CBD20 | GWPCARE1 study (16) | 2.1 (1.0, 4.4) | | | **10.6 (2.3, 48.2) p=0.0022** | 3.1 (0.9, 10.1) p=0.0686 |
| **≥50% Responder rates (Odds ratios, 95%CI) – Full trial population** | | | | | | |
| CBD10 | GWPCARE2 study (17) | 2.2 (1.1, 4.6) | | | **6.8 (1.7, 27.2) p=0.0072** | **11.8 (2.1, 65.4) p=0.0047** |
| CBD20 | GWPCARE2 study (17) | 2.7 (1.3, 5.7) | | | **5.6 (1.4, 22.8) p=0.0174** | **9.6 (1.7, 53.9) p=0.0099** |
| CBD20 | GWPCARE1 study (16) | 2.0 (0.9, 4.3) | 15.0 (4.5, 50.0) | 26.0 (5.5, 123.2) | **7.5 (1.8, 31.5) p=0.0059** | **13.0 (2.3, 74.1) p=0.0039)** |
| Meta-analysis of CBD20 | Gunning et al 2020 (18) | 2.4 (1.4, 4.1) | | | **6.3 (1.7, 23.4) p=0.0065** | **10.8 (2.1, 56.2) p=0.0046** |
| Meta-analysis of CBD10 and CBD20 | | 2.3 (1.5, 3.6) | | | **6.4 (1.8, 23.1) p=0.0045** | **11.1 (2.2, 55.9) 0.0035** |
| **≥50% Responder rates (Odds ratios, 95%CI) – Cannabidiol subgroup taking clobazam** | | | | | | |
| CBD10_CLB | | 2.3 (1.0, 5.7) | | | **6.5 (1.5, 28.8) p=0.0134** | **11.3 (1.9, 67.1) p=0.0076** |
| CBD20_CLB (GWPCARE2) | Gunning et al 2020 (18) | 3.3 (1.3, 8.3) | 15.0 (4.5, 50.0) | 26.0 (5.5, 123.2) | 4.6 (0.99, 20.8) p=0.0508 | **7.9 (1.3, 48.1) p=0.0254** |
| CBD20_CLB (GWPCARE1B) | | 2.9 (1.1, 7.8) | | | **5.2 (1.1, 24.4) p=0.0380** | **9.0 (1.4, 56.3) p=0.0193** |

| Cannabidiol dataset | Source of cannabidiol data | Cannabidiol vs placebo | Fenfluramine 0.7mg/kg/day (max 26mg/day) vs placebo (19) | Fenfluramine 0.4mg/kg/day (max 17mg/day) vs placebo (20) | ITC: Fenfluramine 0.7mg/kg/day vs Cannabidiol | ITC: Fenfluramine 0.4mg/kg/day vs Cannabidiol |
|---|---|---|---|---|---|---|
| Meta-analysis of CBD20_CLB | | 3.1 (1.6, 6.1) | | | **4.8 (1.2, 19.2) p=0.0249** | **8.4 (1.5, 45.6) p=0.0138** |
| Meta-analysis of CBD10_CLB and CBD20_CLB | | 2.8 (1.6, 4.8) | | | **5.4 (1.4, 20.2) p=0.0123** | **9.35 (1.8, 48.5) p=0.0078** |
| **>75% Responder rates (Odds ratios, 95%CI) – Full trial population*** | | | | | | |
| CBD10 | GWPCARE2 study (17) | 6.63 (2.1, 20.7) | 55.1 (6.0, 526.0) | 23.7 (2.9, 191.8) | 8.3 (0.7, 102.3) p=0.0983 | 3.6 (0.3, 38.8) p=0.2953 |
| CBD20 | | 3.3 (1.1, 10.9) | | | **16.6 (1.3, 204.4) p=0.0287** | 7.1 (0.7, 77.6) p=0.1075 |
| CBD20 | GWPCARE1 study (16) | 2.2 (0.8, 6.0) | | | **24.9 (2.2, 287.9) p=0.0100** | **10.7 (1.1, 108.9) p=0.0449** |
| Key: CBD10, cannabidiol 10mg/kg/day; CBD20, cannabidiol 20mg/kg/day; CBD10_CLB, cannabidiol 10mg/kg/day with concomitant clobazam; CBD20_CLB, cannabidiol 20mg/kg/day with concomitant clobazam; ITC, Indirect treatment comparison, 95%CI, 95% confidence interval<br>¶ANCOVA based sensitivity analysis as primary analysis based on median data | | | | | | |

*Table S 9. Adverse events of interest for fenfluramine and cannabidiol licensed dose regimens*

| | **Study 1 (19)** | | **Study 2 (20)** | | **Study 3 (32)** | | **Pooled cannabidiol RCTs – Full trial populations irrespective of concomitant clobazam (18)** | | | **Pooled cannabidiol RCTs - subgroup taking concomitant clobazam (18)** | | |
|---|---|---|---|---|---|---|---|---|---|---|---|---|
| | Placebo (n=40) | FFA0.7 (n=40) | Placebo (n=44) | FFA0.4 (n=43) | Placebo (n=48) | FFA0.7 (n=49) | Pooled Placebo (n=131) | CBD10 (n=72) | Pooled CBD20 (n=139) | Pooled Placebo (n=84) | CBD10_CLB (n=50) | Pooled CBD20_CLB (n=88) |
| **Overall adverse events** | | | | | | | | | | | | |
| At least 1 TEAE | 26 (65) | 38 (95) | 42 (96) | 42 (98) | 40 (83) | 44 (92) | 108 (82) | 61 (85) | 126 (91) | 73 (87) | 44 (88) | 93 (94) |
| Serious TEAE | 4 (10.0) | 5 (12.5) | 7 (15.9) | 6 (14.0) | 2 (4) | 3 (6) | 14 (11) | 15 (21) | 28 (20) | 9 (11) | 11 (22) | 20 (23) |
| Adverse events leading to discontinuation | 0 | 5 (12.5) | 1 (2.3) | 2 (4.7) | 1 (2) | 2 (4) | 1 (1) | (1) | 15 (11) | 1 (1) | 0 | 10 (11) |
| **Adverse events of interest, n (%)** | | | | | | | | | | | | |
| Decreased appetite | 2 (5) | 15 (38) | 5 (11) | 19 (44) | NR | NR | 14 (11) | 12 (17) | 41 (30) | 8 (10) | 9 (18) | 30 (34) |
| Weight decreased | 0 | 2 (5) | NR | NR | NR | NR | 1 (1) | 0 | 7 (5) | 1 (1) | 0 | 5 (6) |
| Somnolence | 3 (8) | 4 (10) | NR | NR | NR | NR | 16 (12) | 19 (26) | 38 (27) | 13 (16) | 17 (34) | 31 (35) |

| | Study 1 (19) | | Study 2 (20) | | Study 3 (32) | | Pooled cannabidiol RCTs – Full trial populations irrespective of concomitant clobazam (18) | | | Pooled cannabidiol RCTs - subgroup taking concomitant clobazam (18) | | |
|---|---|---|---|---|---|---|---|---|---|---|---|---|
| Lethargy | 2 (5) | 7 (18) | 2 (5) | 6 (14) | NR | NR | 5 (4) | 1 (1) | 9 (7) | 5 (6) | 1 (2) | 9 (10) |
| Valvular heart disease | 0 | 0 | 0 | 0 | 0 | 0 | NR | NR | NR | NR | NR | NR |
| Pulmonary arterial hypertension | 0 | 0 | 0 | 0 | 0 | 0 | NR | NR | NR | NR | NR | NR |
| AST increased | NR | NR | NR | NR | NR | NR | 0 | 3 (4) | 11 (8) | 0 | 3 (6) | 10 (11) |
| ALT increased | NR | NR | NR | NR | NR | NR | 0 | 3 (4) | 9 (7) | 0 | 2 (4) | 6 (7) |
| Liver function abnormal | NR | NR | NR | NR | NR | NR | 1 (1) | 0 | 6 (4) | 1 (1) | 0 | 4 (5) |
| Key: ALT, alanine aminotransferase; AST, aspartate aminotransferase; CBD10, cannabidiol 10mg/kg/day; CBD20, cannabidiol 20mg/kg/day in; CBD10_CLB, cannabidiol 10mg/kg/day with concomitant clobazam; CBD20_CLB, cannabidiol 20mg/kg/day with concomitant clobazam; FFA0.4, fenfluramine 0.4mg/kg/day up to a max. 17mg/day; FFA0.7, fenfluramine 0.7mg/kg/day up to a max. 26mg/day; NR, not reported; TEAE, treatment emergent adverse event | | | | | | | | | | | | |

### *Table S 10. Sensitivity analyses around 50% responder rate outcome*

| | **Base case (reference)** | **Sensitivity analyses** | | | | |
|---|---|---|---|---|---|---|
| | **Bayesian NMA Fixed effects** | **Bayesian NMA Fixed effects including Study 3** | **Bayesian NMA Random effects** | **Frequentist NMA Fixed effects** | **Frequentist NMA Random effects** | **Inconsistency (direct vs indirect evidence), p-value*** |
| FFA 0.7 vs Placebo | 15.8 (5.27, 57.2) | 26.4 (11.5, 67.9) | 16.1 (0.81, 340.0) | 14.5 (4.6, 45.8) | 14.5 (4.6, 45.8) | n/a |
| FFA 0.4 vs Placebo | 29.0 (7.10, 222.0) | 29.1 (7.05, 217.0) | 28.7 (1.28, 842.0) | 24.2 (5.2, 112.6) | 24.2 (5.2, 112.6) | n/a |
| CBD 10 vs Placebo | 2.06 (1.04, 4.07) | 2.04 (1.04, 4.05) | 2.05 (0.13, 32.1) | 2.03 (1.0, 4.0) | 2.03 (1.0, 4.0) | 0.56 |
| CBD 20 vs Placebo | 2.39 (1.42, 4.08) | 2.38 (1.40, 4.07) | 2.38 (0.29, 19.2) | 2.36 (1.4, 4.0) | 2.36 (1.4, 4.0) | n/a |
| * The trial network provides direct and indirect evidence only for the comparison of CBD10:CBD20 and CBD10:placebo, both with p-value = 0.56 indicating no significant inconsistencies. | | | | | | |

*Figure S 1. PRISMA flow chart of study screening and identification*

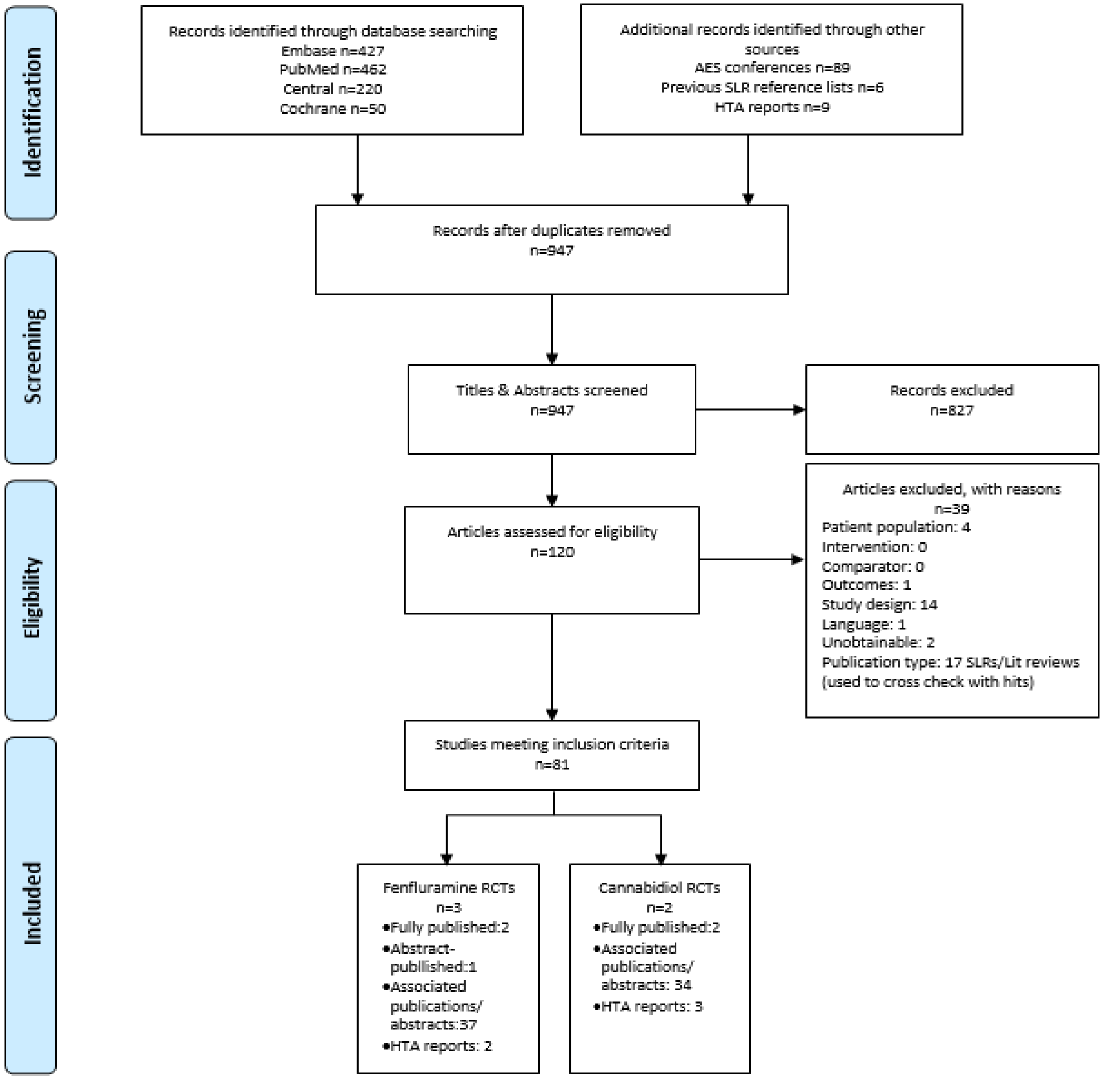

*Figure S 2. Risk of bias assessment using Cochrane Risk of Bias Tool for RCTs (RoB 2)*

Risk of bias domains

| Study | D1 | D2 | D3 | D4 | D5 | Overall |
|---|---|---|---|---|---|---|
| Study 1 (no STP) | + | + | + | + | + | + |
| Study 2 (with STP) | + | + | + | + | + | + |
| Study 3 (no STP) | - | + | + | + | + | - |
| GWPCARE1 (full) | + | + | + | + | + | + |
| GWPCARE2 (full) | + | + | + | + | + | + |
| GWPCARE1 (CLB subgroup) | + | + | + | + | - | - |
| GWPCARE2 (CLB subgroup) | + | + | + | + | - | - |

Domains:
D1: Bias arising from the randomization process.
D2: Bias due to deviations from intended intervention.
D3: Bias due to missing outcome data.
D4: Bias in measurement of the outcome.
D5: Bias in selection of the reported result.

Judgement
- Some concerns
+ Low

*Figure S 3. NMA of mean placebo-adjusted monthly reduction from baseline in MCSF (on log scale)*

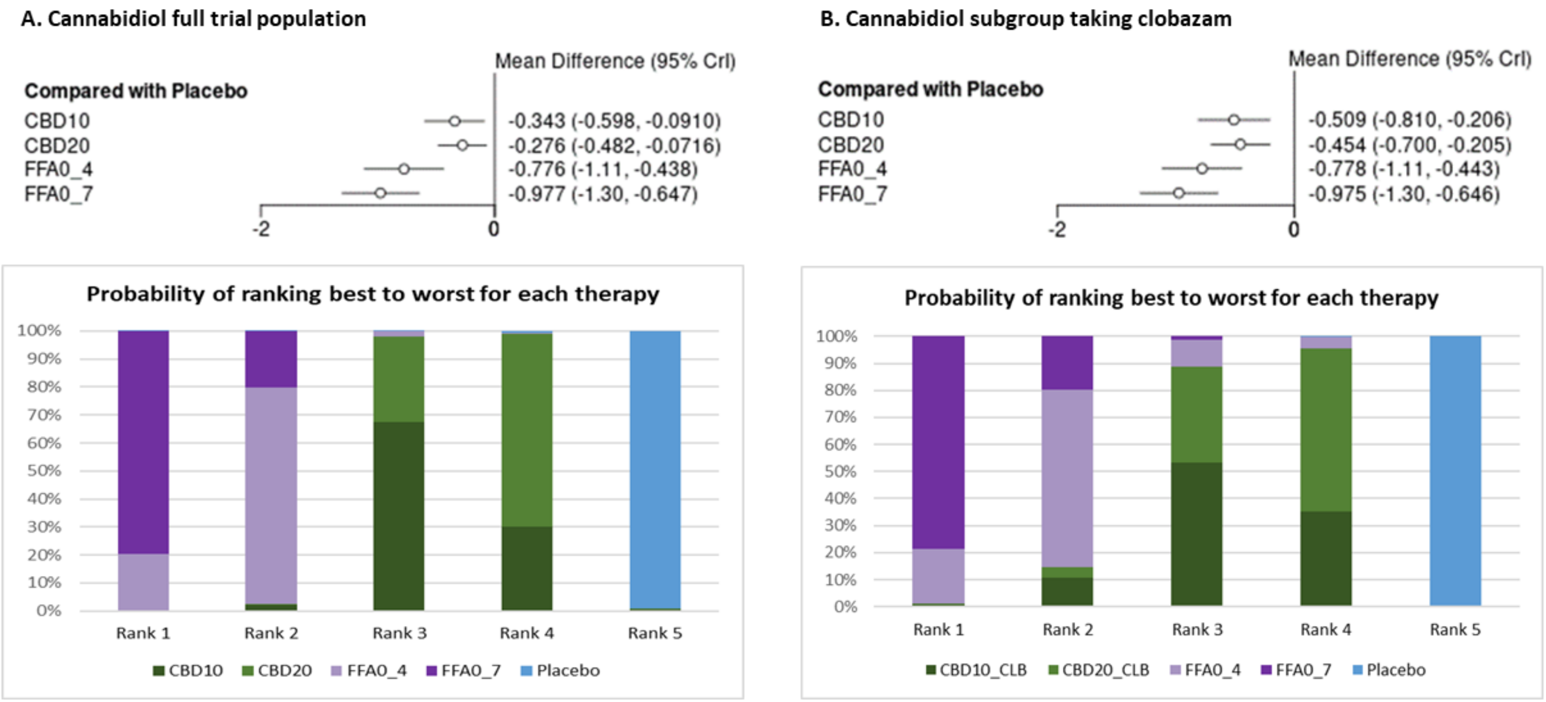


Key: 95%CrI, 95% credible interval; CBD10, cannabidiol 10mg/kg/day; CBD10_CLB, cannabidiol 10mg/kg/day plus clobazam; CBD20, cannabidiol 20mg/kg/day; CBD20_CLB, cannabidiol 20mg/kg/day plus clobazam; CSF, convulsive seizure frequency; FFA0_4, fenfluramine 0.4mg/kg/day up to a max 17mg/day; FFA0_7, fenfluramine 0.7mg/kg/day up to a max 26mg/day. Rank 1 is most effective therapy, Rank 5 least effective therapy

*Figure S 4. Incidence of 100% reduction in MCSF (fully published trials)*

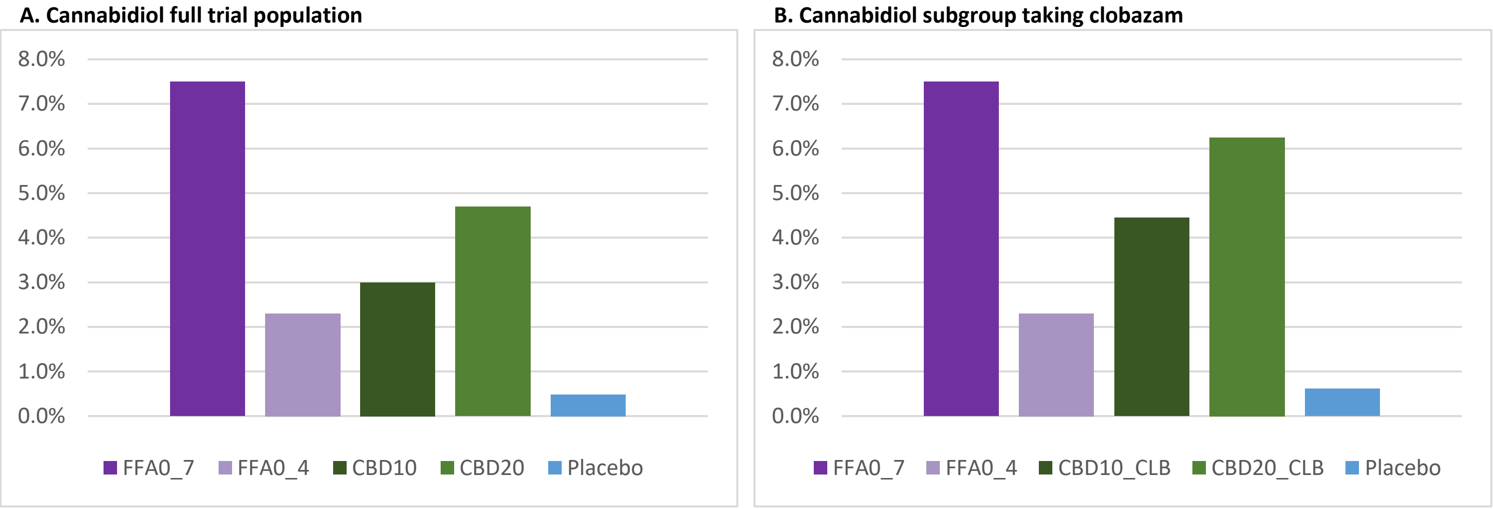


Key: CBD10, cannabidiol 10mg/kg/day; CBD10_CLB, cannabidiol 10mg/kg/day plus clobazam; CBD20, cannabidiol 20mg/kg/day; CBD20_CLB, cannabidiol 20mg/kg/day plus clobazam; FFA0_4, fenfluramine 0.4mg/kg/day up to a max 17mg/day; FFA0_7, fenfluramine 0.7mg/kg/day up to a max 26mg/day; MCSF, monthly convulsive seizure frequency.

*Figure S5. Best-worst probability ranking for incidence of serious TEAEs*

**A. Cannabidiol full trial population**

**B. Cannabidiol subgroup taking clobazam**

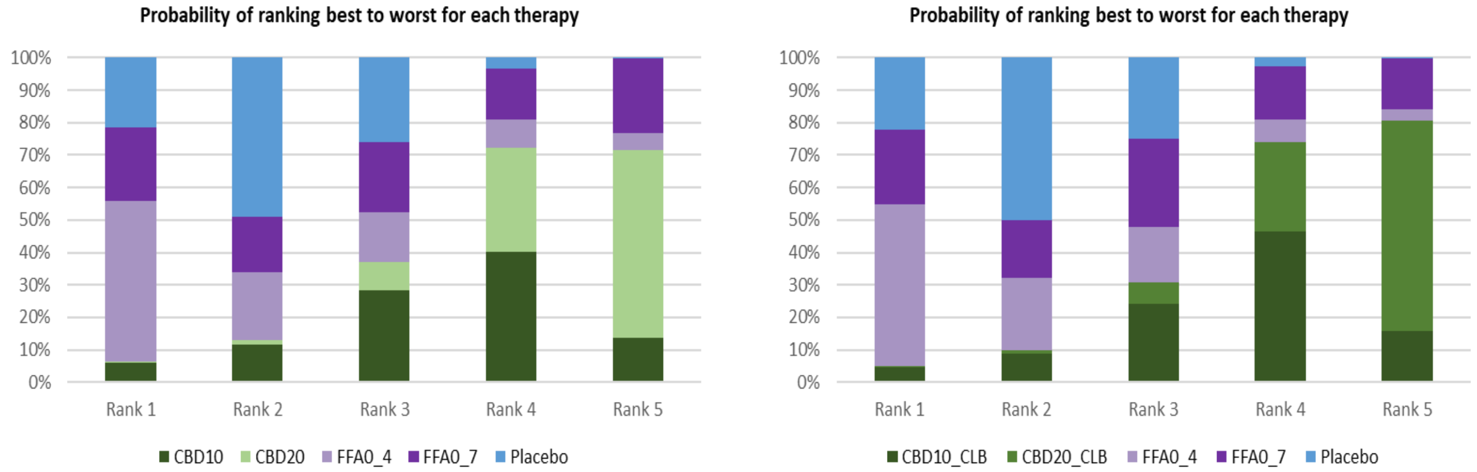


Key: CBD10, cannabidiol 10mg/kg/day; CBD10_CLB, cannabidiol 10mg/kg/day plus clobazam; CBD20, cannabidiol 20mg/kg/day; CBD20_CLB, cannabidiol 20mg/kg/day plus clobazam; FFA0_4, fenfluramine 0.4mg/kg/day up to a max 17mg/day; FFA0_7, fenfluramine 0.7mg/kg/day up to a max 26mg/day; MCSF, monthly convulsive seizure frequency. Rank 1 (best) has lowest odds of serious TEAEs, Rank 5 (worst) has highest odds of serious TEAEs
